# Neglected Tropical Diseases in conflict-related humanitarian emergency settings: a systematic review of the literature

**DOI:** 10.1101/2025.08.15.25333750

**Authors:** Charlotte Bestwick, Xavier Badia-Rius, Sergio Lopes, Louise A. Kelly-Hope, Emma M. Harding-Esch

## Abstract

**Background:** Neglected Tropical Diseases (NTDs) are a group of 21 conditions affecting an estimated one billion people worldwide, causing significant mortality and morbidity. The World Health Organization (WHO) has targeted their control, elimination or eradication by 2030, whilst highlighting that conflict and humanitarian emergencies are risks to achieving this.

**Methods:** A systematic review of peer-reviewed literature was undertaken, using terms related to conflict-related humanitarian emergencies (CRHE), based on the Armed Conflict Location & Event Data Project codebook terms for defining emergencies, and terms including all WHO-defined NTDs. After a two-stage screening process, 26 papers were included.

**Results:** Common challenges for NTD programmes in CRHE were healthcare access, infrastructure, population movement and data quality. Multi-sectorial collaboration between actors in conflict-settings was offered as a learning for NTD programmes, along with community participation and decentralisation. Evidence gaps were identified in the knowledge base for certain NTDs and in high-quality evidence for interventions in CRHE.

**Conclusions:** Collaboration is a key area for focused improvement for NTD programmes in CRHE. This should be across sectors, and extend to research and analysis approaches, to ensure the most effective interventions are identified and implemented, and NTD road map targets can be achieved.

## Introduction

Neglected Tropical Diseases (NTDs) are a broad range of 21 conditions, affecting an estimated one billion people worldwide, often those living in poverty (1). NTDs contribute to an estimated 200,000 deaths annually, with additional morbidity and many NTD survivors enduring economic and social hardship and stigma (2).

The World Health Organization (WHO) published a road map for NTD control, elimination and eradication in 2012, which initially included targets to be achieved by 2020 (3). These were subsequently expanded in the road map for NTDs 2021–2030 (4), to relate closer to the United Nation’s (UN) Sustainable Development Goals (5), and with the date for completion extended to 2030. The 2021-2030 road map highlights the need to strengthen health systems against humanitarian emergencies such as conflict, to improve resilience and help overcome challenges to achieving the targets (4). The annual WHO Global Report on NTDs in 2024 voiced this more clearly, stating conflict and humanitarian crises exacerbate the risk of NTDs in affected populations and negatively affect, indeed sometimes prohibit, actions to progress NTD programme implementation (1). Whilst identifying this as a problem, little evidence have been collated on this issue, and at present there are no formal guidelines for NTD programmes in conflict settings, aside from the 2025 preferred practices developed by the International Coalition for Trachoma Control (6).

Conflict is recognised by the UN as one of three drivers of humanitarian emergencies, alongside climate and economic factors (7). The mechanism for the association between conflict-related humanitarian emergencies (CRHE) and NTD programmes is complex and multi-faceted (8). CRHE settings often involve disruption of “normal life”, including: population displacement (often with the creation of camps to house displaced people); lack of water and sanitation; and interruption to healthcare, including access to facilities and medications. This can leave populations vulnerable to poverty (or exacerbate existing poverty) and disease-spread, and can lead to stalling of NTD control efforts, or even re-introduction of disease (4, 9). There are numerous specific examples of the association between CRHE and NTDs historically, including evidence of a demonstrable dose-response link between increasing levels of conflict and terror with the incidence of leishmaniasis, with calls for leishmania programmes to ensure the impact of conflict and terror on their populations are accounted for when planning interventions (10).

Whilst there is a growing body of evidence to demonstrate the challenges CRHE pose to NTD control programmes, specific guidance for NTD programmes to negotiate, and ultimately overcome, them are yet to be produced. To enable shared learnings between programmes and support future practice and policy by generating evidence-based recommendations, we conducted a systematic review to establish existing examples of challenges, strategies, solutions and experiences NTD programmes have encountered and implemented in CRHE, and to identify gaps in evidence.

## Materials and methods

### Study protocol and registration

The study protocol was registered on PROSPERO (CRD42024554100) (11). The search findings were reported following Preferred Reporting Items for Systematic reviews and Meta-Analyses (PRISMA) guidelines (12).

### Search Strategy

A systematic review of peer-reviewed literature using electronic databases – Medline, Embase and Global Health – to identify relevant literature for this project was undertaken. These databases were selected to reflect the global focus of this review, ensuring inclusion of literature from wide geographic regions. The search terms were developed from two main components joined together with AND: terms related to conflict-related humanitarian emergencies (based on Armed Conflict Location & Event Data Project [ACLED] Conflict Index Codebook terms (13)), and terms related to NTDs (including all conditions covered by the WHO definition of NTDs (1)). ACLED is a publicly-available project beginning in 2005, which collates and maps data on all global reports of conflict events (14, 15). The initial search was constructed in Ovid Medline, with each search concept using free-text, and MeSH terms wherever possible. Two filters were applied to results: English language, and date range of 1^st^ January 2005 to the search date (30^th^ May 2024). This search structure was then transferred to the two other databases. The search strategy is available in Supplementary Table 1.

### Eligibility criteria

All papers conducting analysis or reporting of primary data on human study populations relating to NTD programmes (at least one WHO-defined NTD (1)) in CRHE settings (meeting ACLED codebook criteria), written in English, from 1^st^ January 2005 to 30^th^ May 2024 were included. Papers were excluded if they were missing abstract or full-text, involved secondary analysis (including mathematical modelling, opinion or systematic reviews), or focused on non-human models or data. Grey literature was excluded due to the large volume of results from the peer-reviewed database search. Full eligibility criteria are in Supplementary Table 2.

### Study screening and selection

The database search output was imported into EndNote, with duplicates removed and a preliminary screen to remove posters, editorials and opinion pieces. All retained results were then reviewed in a two-step process. First, titles and abstracts were reviewed by two independent reviewers, with the primary reason for exclusion recorded in Microsoft Excel. Any studies that fulfilled the inclusion criteria or where there was disagreement between the reviewers were included in the next stage. After this stage, any studies where the full-text could not be located were excluded. In the second stage of screening, one reviewer screened all results, with a second reviewer screening a randomly-selected 15% subset of papers. Disputed studies for inclusion between the two reviewers were discussed between the study group to reach a consensus agreement. Reasons for exclusion at the full-text review stage were recorded in Excel.

### Data extraction

Data extraction was performed in Excel, with headings including: population, CRHE detail, study design, NTD(s) in focus, challenges encountered by NTD programme, solutions to overcome the challenges, and gaps in evidence identified. The data extraction table is summarised in Supplementary Table 3. A second reviewer performed data extraction on a random 15% subset of papers, with their output compared to the first reviewer, to ensure consistency.

### Data analysis

The study selection process and characteristics were recorded in Excel and presented as per PRISMA guidelines (12) using R (16) (see Figure 1). The extracted data were analysed thematically (17) to identify commonly-recurring challenges and solutions of NTD programmes in CRHE settings, and gaps in evidence.

**Figure 1.**
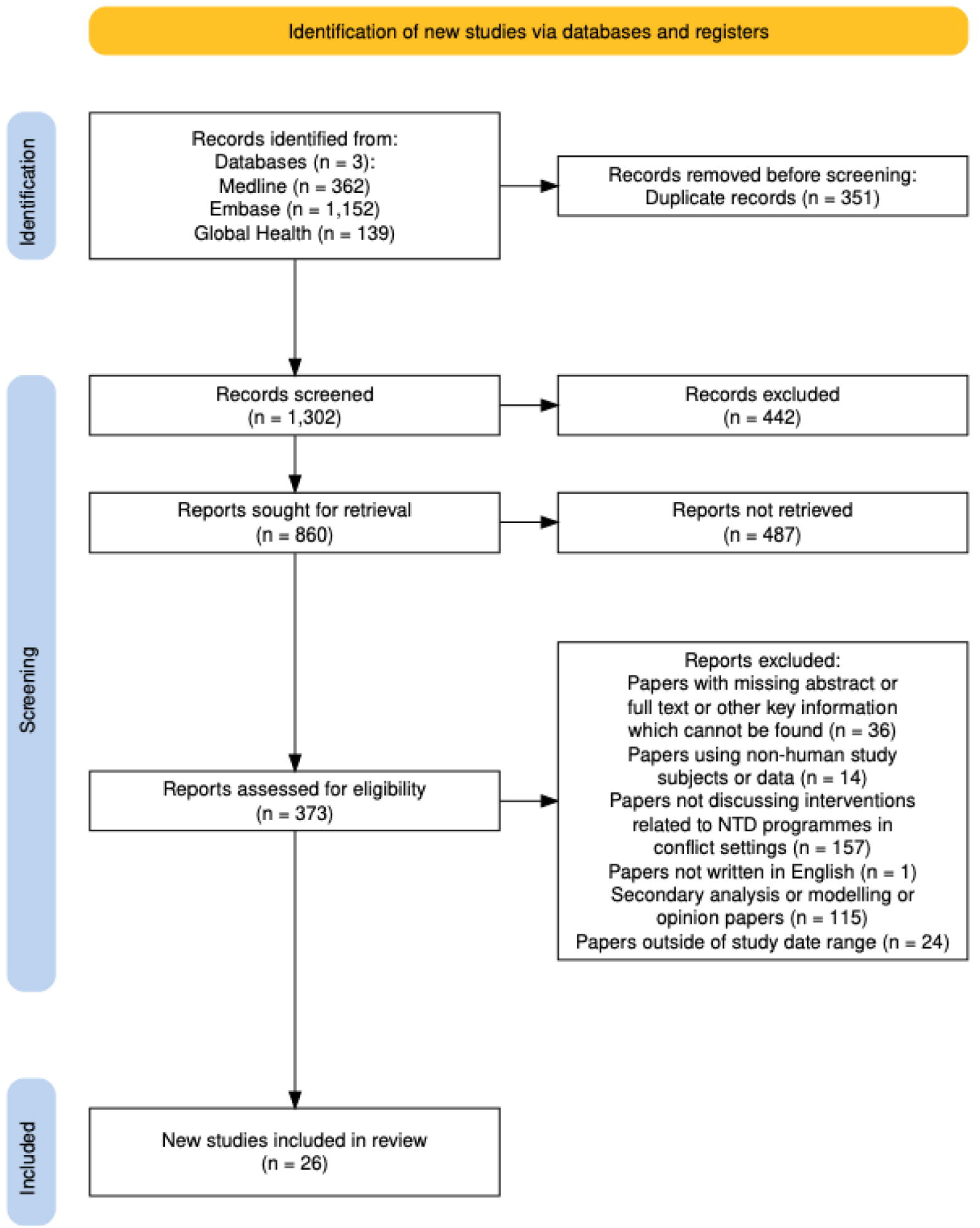
PRISMA Flowchart illustrating how the 26 studies were selected for inclusion

### Quality Assessment

Each included study underwent a risk of bias assessment. Critical appraisal tools produced by the Joanna Briggs Institute (JBI) were used, in the form of specific checklists for each study type (18). JBI checklists were chosen due to the wide range of study designs with corresponding checklists. Where a dedicated JBI checklist for the study design did not exist, the most appropriate checklist was completed. The outcome of the checklist for each study was scored: the numerator of the number of “yes” response questions over the denominator of the total number of applicable questions from that checklist was converted into a numerical value, with 1 being the maximum and 0 the minimum score for quality. JBI checklists are summarised in Supplementary File 4.

## Results

### Study searching and selection

Of the 1653 results combined from the three database searches, 1302 remained after removal of 351 duplicates. These references were then screened and opinion articles (n=442) removed prior to title/abstract screening. Of the remaining 860 articles for screening, 373 were found to be eligible for second stage screening. Papers where full-texts could not be sourced (n= 36) were removed, leaving 338 papers for review. Following application of the eligibility criteria to full texts, a final total of 26 papers were included (Figure 1).

### Study characteristics and coverage

Of the 26 included papers, one was an interventional randomised controlled trial (RCT), and the remaining were observational studies. The publication dates were broadly equally spread across the time-period boundaries of this review, from 2006 to 2024. All 26 papers collected data from countries in Africa or Asia, with most studies taking place in Sri Lanka, Turkey and Syria (4, 3 and 3, respectively) and none from the Americas (Figure 2). The papers described a range of CRHE settings; several were conducted by NTD programmes operating within active-conflicts (19–30), and others in conflict-adjacent regions encountering people displaced due to a conflict (31–41). One study reviewed 32 communicable diseases in one country (Iraq), which included numerous NTDs in its scope. Across the 25 remaining studies, there were 9 NTDs in focus, with the most studied NTD being cutaneous leishmaniasis (8 papers). A summary of the papers is in Supplementary Table 3.

**Figure 2.**
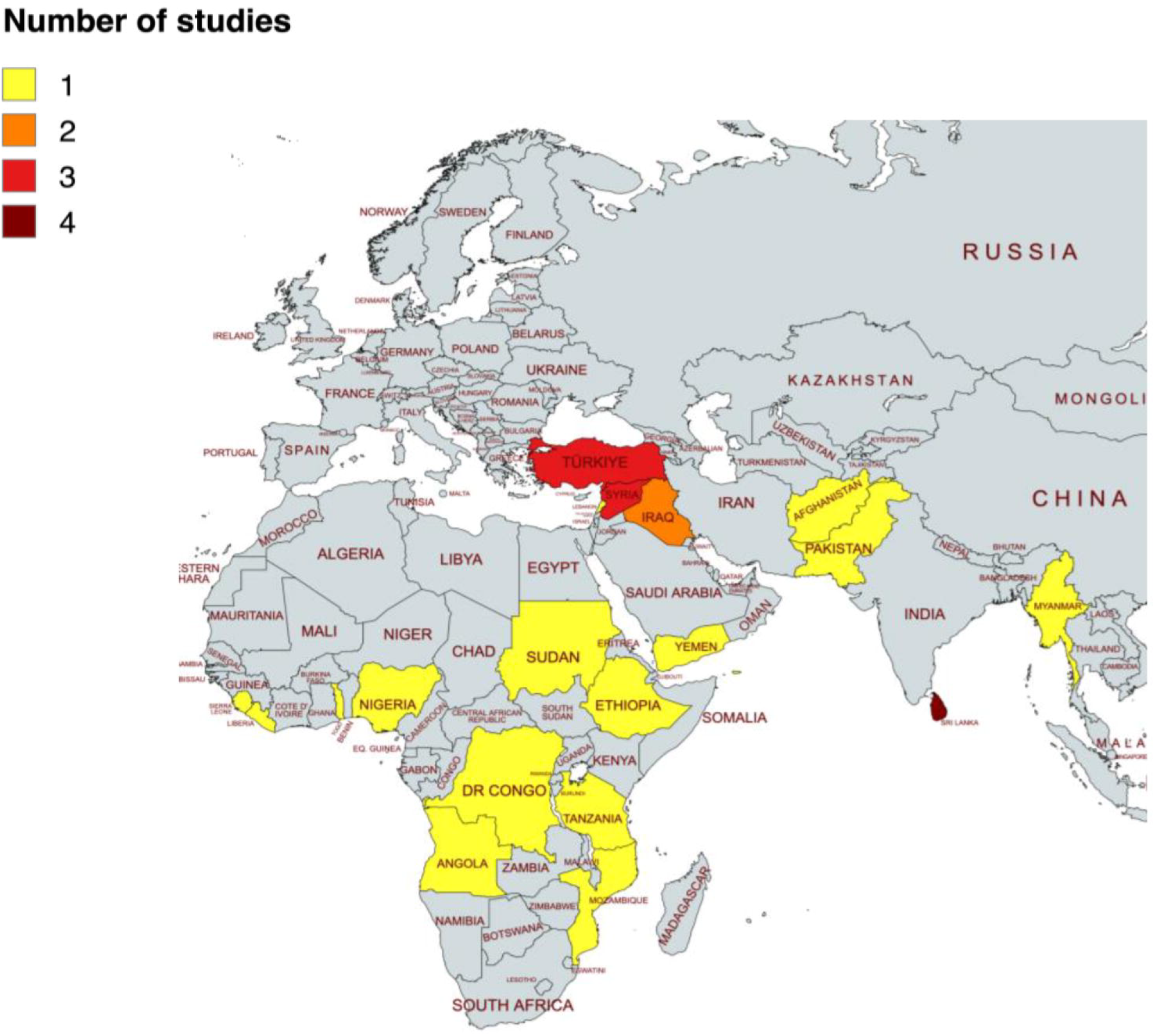
Geographic spread of the 26 papers in this review, created using mapchart.net. The American Regions are excluded from this figure as no studies were included from that region.

### Quality of the evidence

Most studies (19 of 26) scored highly on JBI checklists, indicating they were of high quality and low risk of bias (Supplementary File 4). Nine studies had incomplete reporting of participant clinical and demographic information, but this was often reflective of a true evidence gap, due to poor-quality health records (19, 27, 28, 30, 42), weak surveillance systems (21) or limited resources to complete thorough investigations and follow-up (20, 39, 43). Only one study was an interventional RCT, and this scored poorly on quality assessment (with problematic methodology including lack of blinding and not using intention-to-treat analysis methods) (24).

### Thematic synthesis of results

A summary of the challenges encountered by the NTD programmes reported in the studies, together with the solutions, is shown in Figure 3.

**Figure 3.**
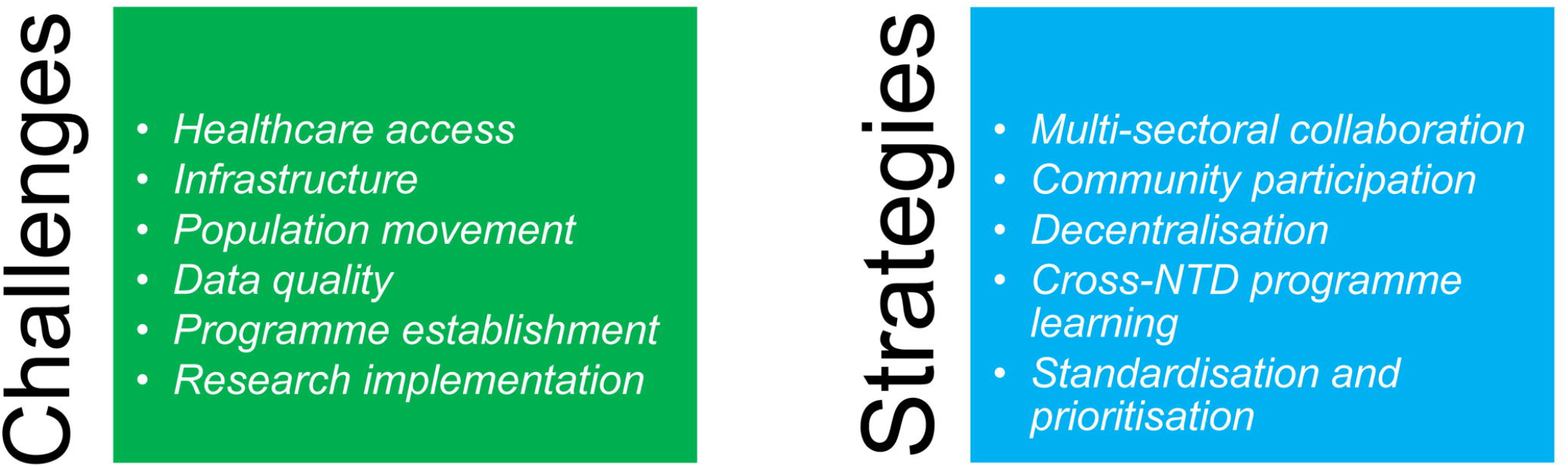
Summary of the themes arising from the 26 included papers, separated by challenges encountered by Neglected Tropical Disease (NTD) programmes in conflict-related humanitarian emergency settings, and strategies or learnings to overcome these challenges.

### Challenges encountered by NTD programmes

#### Healthcare access

The most common challenge was the impact of conflict on healthcare access. Reasons for this included: physical destruction of facilities during conflict (26, 34), disruption to drug and equipment supply-chains (22, 28–30), workforce shortages due to high security risk and little capacity to train staff (26–28, 42), and deterioration of functioning healthcare systems (19, 20, 24, 30, 32). Interruption to programmes for vaccination (29), surveillance (21), case-finding (23), awareness (22), and mapping surveys (25) led to limited capacity for disease control, little knowledge of disease epidemiology, and poor outcomes for those affected.

#### Infrastructure

The impact of conflict on wider infrastructure limited programmes, for example areas becoming inaccessible through the destruction or closure of roads (19, 22). The breakdown of municipal services resulting in accumulation of rubbish in which vectors can nest (19, 22) or bomb-damage to buildings in which vectors could live (26), resulted in NTD vector breeding sites being created during conflict, facilitating their multiplication and increasing risk for populations. Conflict was also seen to generally degrade living conditions, with multiple accounts of widespread famine, increasing susceptibility to infections (21, 22).

#### Population movement

Mass population movement was a common challenge for NTD programmes, with changing disease epidemiology when displaced people moved from a highly-endemic region to a low- or non-endemic region during a time of conflict (19–21, 27, 30, 31, 36–39). This, coupled with healthcare and surveillance system breakdown, led to undetected disease-spread and unprepared healthcare resources (38, 43). Unfamiliarity with a new country or region’s healthcare system led to diagnostic delay and poor health-seeking behaviour (38), and disruption to drug administration programmes (24, 25). Population movement was not always via a documented route, with some people left unaccounted for, and thus not included in health systems programming (38, 40). Movement was often seen into camps for displaced people or refugees, which were frequently in hard-to-reach, rural areas (33), with poor conditions, overcrowding, and limited appropriate water, sanitation and hygiene (WASH) access, all of which increase disease risk (25, 27, 29, 31–35, 38, 40). Population movement not related to camps included displacement away from fighting, resulting in an influx into cities, which then also became congested and overcrowded, increasing infection risk (30).

#### Data quality

Accurate data was a challenge for NTD programmes in regions with multiple actors at play, including multiple parallel systems for case reporting for the same condition in Sri Lanka (42), and conflicting case definitions being used by international organisations operating alongside one another in Syria, leading to unreliable case reports (26). Reporting inconsistencies (21, 38), poor medical registries, population demographics, or statistics, were also cited in multiple studies (26, 30). Poor or limited data impaired NTD programmes’ ability to make informed programmatic decisions.

#### Programme establishment

Several papers discussed the difficulties of establishing a programme in CRHE settings if no previous programme existed. Challenges reported included little experience by healthcare professionals or health systems of managing NTDs (38, 43), poor diagnostic capacity (22, 42), inadequate vector-control methods (20), and lack of centralised reporting or surveillance systems (43). Starting from a poor baseline meant NTD programmes could lack experienced guidance and may require greater resources to become fully operational.

#### Research implementation

Multiple studies reported problems in obtaining resources or funding (19, 20, 36), and being physically unable to access high-burden regions as these were frequently rural or poor, with limited healthcare coverage (23, 38). The difficulties of conducting NTD research in CRHE settings posed a challenge to build a body of evidence-based research.

### Solutions, Strategies and Learnings of NTD programmes

#### Multi-sectoral collaboration

Multiple studies stated that the crux of successful NTD programming in CRHE settings is multi-sectoral collaboration, which for some programmes may only occur once political stability is achieved (21, 23, 26). This collaboration involves local and national governments, international aid agencies (25, 38), and existing control programmes for other infectious diseases (27, 32). How to go about collaboration did vary by setting but was considered critical across the board. Multiple studies on cutaneous leishmaniasis agreed that multisectoral activities involving education, research, and innovation were required for disease-control in CRHE settings (19–21).

#### Community participation

Calls for community participation were made in several studies, with use of education and awareness to encourage health-seeking behaviour (19, 22, 28, 38), and increased use of community health workers (41, 44), particularly in poorly-resourced healthcare systems (43), to enable appropriate NTD management. In instances of population displacement, several papers suggested focusing surveillance and control initiatives (including WASH and targeted education campaigns) in refugee camp-settings to see greatest impact on minimising disease spread (27, 30, 33–35, 39, 40, 42).

#### Decentralisation

Using reporting networks with congruent case definitions and clinical diagnoses by trained community practitioners were highlighted as key recommendations for CRHE settings (26,42,43). Prioritising the quick restoration of primary healthcare after conflict was also shown to allow health systems to rapidly resume, and even resulted in a declining trend for some zoonotic NTDs, such as brucellosis (29).

#### Cross-NTD programme learning

Solutions offered may be derived from a particular NTD, but their learnings are adaptable for sharing more widely in the NTD community. Examples include the identification of high-risk groups, and prioritising the scale-up of interventions to target these groups to prevent trachoma infection (34).

#### Standardisation and prioritisation

Standardisation of therapy and consistent access to essential medicines should be adopted by programmes in active conflict settings, as demonstrated by the reduction in snake bite morbidity and mortality during times of consistent antivenom and adrenaline supply (28).

### Gaps in Evidence

The lack of, or significant problems with, population statistics were identified by several papers, with patchy coverage of surveillance systems, unreliable clinical records and inconsistent demographic detail proving a problem for programmatic decision-making (20,26,29,30). Specific diseases were found to be poorly researched (19,27), or have limited treatment options available, notably cutaneous leishmaniasis (19,38) and soil-transmitted helminths (32). Several studies also detailed the lack of proper monitoring and evaluation systems for NTD programmes in CRHE settings, with absence of surveillance, post-validation surveys or feedback (25,36,37,40).

## Discussion

This review found that in CRHE settings, poor healthcare access, disruption to existing programmes, mass population movement and degradation of living conditions are key challenges for NTD programmes. Solutions to these challenges included multi-sectoral collaboration, community participation, and identification of high-risk population groups for targeted interventions. These principal findings are in-line with several of the UN Sustainable Development Goals (5), including SDG 3 (good health and wellbeing), SDG 16 (peace and strong institutions) and SDG 17 (partnerships) (45).

CRHE are inherently challenging environments to implement any healthcare programme, often with a complete breakdown in health and wider infrastructure. Limited funding and resources constrict research capacity in these situations, demonstrated by the small number of high-quality RCT studies to guide evidence-based medicine (46). This paper adds to the call for more funding and research into this field (47,48), especially in relation to the challenges raised such as improving healthcare access to rural areas (23,38).

This study showed that the prioritisation of data collection during and after a conflict can prove transformative (20,25,29,43). NTD programmes require ongoing data for monitoring and evaluation, even after achieving NTD road map targets, and especially in regions where conflict may arise, which could threaten the collapse of control programmes, and risk disease re-introduction. The third “pillar” of the NTD road map is to ‘facilitate country ownership’(4), hence the importance of country programmes to have systems in place to monitor potentially vulnerable areas, and be held accountable by annually reporting their findings (1).

Shared learnings offered to policymakers to achieve better data in CRHE included targeting surveillance efforts in camps for displaced populations to efficiently contain outbreaks (27,30,33–35,39,40,42), or innovative ideas such as using public ‘smart cards’ to store health records (29). There are also existing data sources that can contribute to population-level data; it has been shown that routinely reported data from mass drug administration NTD programmes (such as numbers of drugs distributed and national census data) can be adequately valid to report programme coverage (the proportion of people in need of treatment who actually receive a dose)(49). Implementing new ideas whilst continuing to develop existing methods can enhance data accuracy, strengthen data quality and ultimately lead to more effective decision-making for NTD programmes.

Multi-sectoral collaboration is a key strategy highlighted in this review. This may involve community involvement in strategy development, collaboration between individual disease programmes, and/or collaboration between all actors at play in CRHE. The need for co-ordinated approaches is emphasised in the NTD road map (4), and WHO or other organisations, such as the NTD NGO Network, appear to have a key role in co-ordination to address the challenges of operating in CRHE. Collaboration also extends to the collection of data, with suggestions to use stratified disease assessment methods that account for displacement, conflict and geographic access when planning interventions (25). One example of a collaborative innovation already in use is the Conflict Exposure Calculator, created from a partnership between ACLED and WorldPop (50).

Many of the included studies in this review scored highly on quality assessment, indicating trustworthy and relevant results. Nevertheless, several studies had incomplete reporting of study participants’ clinical or demographic data (19,27,28,30,42), which was found to reflect a true evidence gap, contributing to the learnings from this review. The observational approach of many of the papers included may reflect the complexities of conducting rigorous scientific study in the challenging environment of CRHE. This is further compounded by the fact that NTDs, by definition, are relatively neglected by research (51). Therefore, it is not unexpected that this review identified large evidence gaps for NTD programmes in CRHE, such as weak evidence or experience of WASH interventions being implemented during humanitarian crises for a trachoma programme (40).

Whilst the reason many NTDs were not covered in this review may be due to a lack of studies on these diseases (as found in other systematic reviews (48)), it could also be due to limits applied to the database searches (English language, publication date 2005-2024, human population). The limit of English language may explain the geographic spread of results, and why regions known to have high NTD prevalences, such as South America and the Caribbean (52), were excluded. Several NTDs are zoonotic, such as rabies, so excluding non-human study populations perhaps explains why there were no papers on rabies in the final set of studies. A further limitation is not including grey literature resources, which may have enhanced the understanding of NTD programmes’ experience in CRHE.

This review focused on conflict as the cause of humanitarian emergencies, but it is important to note there are other causes, such as climate and economic factors, including extreme weather events, population displacement and food insecurity (7). Novel approaches that take a multi-sectoral view of the complex interactions between conflict, climate and non-conflict related population displacement as risks to NTD programmes are now being used to stratify risk by some research groups (25,53). This multi-faceted collaborative approach should be encouraged to ensure all these challenges that threaten NTD programmes are suitably quantified, so strategies to overcome them can be implemented, and NTD road map targets can be achieved.

## Conclusions

This systematic review identified key challenges for NTD programmes in CRHE, including poor data, infrastructure destruction, and population movement. Limited high-quality trials evaluating interventions and absence of clinical knowledge in some disease areas mean gaps in evidence remain. A key area for focused improvements for NTD programmes in CRHE is collaboration. This collaboration should be across sectors, including communities, local and national governments, aid organisations, different disease programmes, and extend to research and analysis approaches, to ensure the most effective interventions are identified and implemented.

## Data Availability

All relevant data are within the manuscript and its Supporting Information files.

## Authors statements

### Authors’ contributions

LAKH and EMHE conceived the study. CB, LAKH and EMHE designed the study protocol. CB, XBR and SL collected the data. CB analysed the data. CB, LAKH and EMHE interpreted the data. CB, LAKH and EMHE drafted the manuscript; XBR and SL critically revised the manuscript. All authors read and approved the final version. EMHE is the guarantor of the paper.

### Funding

No specific funding supported this work.

### Competing interests

EMHE receives salary support from the International Trachoma Initiative, which receives an operating budget and research funding from Pfizer Inc., the manufacturers of Zithromax® (azithromycin).

### Ethical approval

The London School of Hygiene & Tropical Medicine Research Governance & Integrity Office confirmed that ethical approval was not required (ref: 30756).

## Notes

### Funding Statement

This study did not receive any funding.

