## Supplementary Files for "Neglected Tropical Diseases in conflict-related humanitarian emergency settings: a systematic review of the literature"

**Supplementary file 1: Medline database search terms**

| **#** | **Query** | **Results from 30 May 2024** |
| --- | --- | --- |
| 1 | exp Buruli Ulcer/ | 736 |
| 2 | exp Mycobacterium ulcerans/ | 911 |
| 3 | "buruli ulcer".map. [mp=title, book title, abstract, original title, name of substance word, subject heading word, floating sub-heading word, keyword heading word, organism supplementary concept word, protocol supplementary concept word, rare disease supplementary concept word, unique identifier, synonyms, population supplementary concept word, anatomy supplementary concept word] | 1,236 |
| 4 | 1 or 2 or 3 | 1,397 |
| 5 | "Chagas disease".mp. or exp Chagas Disease/ | 19,300 |
| 6 | "trypanosoma cruzi".mp. or exp Trypanosoma cruzi/ | 17,982 |
| 7 | 5 or 6 | 25,411 |
| 8 | exp Dengue/ | 16,717 |
| 9 | Dengue.mp. [mp=title, book title, abstract, original title, name of substance word, subject heading word, floating sub-heading word, keyword heading word, organism supplementary concept word, protocol supplementary concept word, rare disease supplementary concept word, unique identifier, synonyms, population supplementary concept word, anatomy supplementary concept word] | 30,640 |
| 10 | exp Chikungunya virus/ or exp Chikungunya Fever/ or chikungunya.mp. | 7,919 |
| 11 | 8 or 9 or 10 | 34,799 |
| 12 | dracunculiasis.mp. or exp Dracunculiasis/ | 992 |
| 13 | exp Dracunculus Nematode/ | 267 |
| 14 | "guinea worm".mp. | 477 |
| 15 | 12 or 13 or 14 | 1,152 |
| 16 | echinococcosis.mp. or exp Echinococcosis/ | 22,649 |
| 17 | clonorchiasis.mp. or exp Clonorchiasis/ | 1,281 |
| 18 | clonorchis.mp. | 1,609 |
| 19 | opisthorchiasis.mp. or exp Opisthorchiasis/ | 1,933 |
| 20 | opisthorchis.mp. or exp Opisthorchis/ | 1,918 |
| 21 | exp Fasciola/ or fasciola.mp. | 6,410 |
| 22 | fascioliasis.mp. or exp Fascioliasis/ | 4,706 |
| 23 | paragonimiasis.mp. or exp Paragonimiasis/ | 1,516 |
| 24 | paragonimus.mp. or exp Paragonimus/ | 1,427 |
| 25 | "food borne trematodes".mp. | 40 |
| 26 | 17 or 18 or 19 or 20 or 21 or 22 or 23 or 24 or 25 | 13,212 |
| 27 | exp trypanosoma brucei brucei/ or exp trypanosoma brucei gambiense/ or exp trypanosoma brucei rhodesiense/ | 9,596 |
| 28 | exp Trypanosomiasis, African/ or "human african trypanosomiasis".mp. | 6,817 |
| 29 | "sleeping sickness".mp. | 2,915 |
| 30 | 27 or 28 or 29 | 13,832 |
| 31 | leishmaniasis.mp. or exp Leishmaniasis/ | 33,052 |
| 32 | exp Leishmania/ or leishmania.mp. | 32,139 |
| 33 | 31 or 32 | 43,532 |
| 34 | leprosy.mp. or exp Leprosy/ | 27,632 |
| 35 | "mycobacterium leprae".mp. or exp Mycobacterium leprae/ | 7,556 |
| 36 | 34 or 35 | 29,220 |
| 37 | "lymphatic filariasis".mp. or exp Elephantiasis, Filarial/ | 4,391 |
| 38 | exp Elephantiasis, Filarial/ or elephantiasis.mp. | 4,576 |
| 39 | 37 or 38 | 5,960 |
| 40 | Mycetoma.mp. or exp Mycetoma/ | 2,868 |
| 41 | chromoblastomycosis.mp. or exp Chromoblastomycosis/ | 1,316 |
| 42 | sporotrichosis.mp. or exp Sporotrichosis/ | 2,586 |
| 43 | "deep mycoses".mp. | 296 |
| 44 | 40 or 41 or 42 or 43 | 6,738 |
| 45 | exp Noma/ or noma.mp. | 967 |
| 46 | onchocerciasis.mp. or exp Onchocerciasis/ | 5,384 |
| 47 | onchocerca.mp. or exp Onchocerca/ | 3,318 |
| 48 | "river blindness".mp. or exp Onchocerciasis, Ocular/ | 747 |
| 49 | 46 or 47 or 48 | 6,309 |
| 50 | rabies.mp. or exp Rabies/ | 17,746 |
| 51 | scabies.mp. or exp Scabies/ | 5,522 |
| 52 | tungiasis.mp. or exp Tungiasis/ | 384 |
| 53 | exp Ectoparasitic Infestations/ or ectoparasitoses.mp. | 23,163 |
| 54 | 51 or 52 or 53 | 24,747 |
| 55 | schistosomiasis.mp. or exp Schistosomiasis/ | 29,558 |
| 56 | schistosoma.mp. or exp Schistosoma/ | 24,919 |
| 57 | 55 or 56 | 36,928 |
| 58 | "soil transmitted helminths".mp. | 1,324 |
| 59 | roundworm.mp. | 1,088 |
| 60 | "ascaris lumbricoides".mp. or exp Ascariasis/ or exp Ascaris lumbricoides/ | 8,427 |
| 61 | whipworm.mp. | 451 |
| 62 | "trichuris trichiura".mp. or exp Trichuris/ | 3,096 |
| 63 | hookworm.mp. | 6,012 |
| 64 | "necator americanus".mp. or exp Necatoriasis/ or exp Necator americanus/ | 1,067 |
| 65 | "ancylostoma duodenale".mp. or exp Ancylostoma/ | 1,577 |
| 66 | "strongyloides stercoralis".mp. or exp Strongyloidiasis/ or exp Strongyloides stercoralis/ | 5,262 |
| 67 | 58 or 59 or 60 or 61 or 62 or 63 or 64 or 65 or 66 | 20,819 |
| 68 | "snake bites".mp. or exp Snake Bites/ | 6,089 |
| 69 | "snakebite envenoming".mp. | 428 |
| 70 | 68 or 69 | 6,247 |
| 71 | taeniasis.mp. or exp Taeniasis/ | 8,495 |
| 72 | "taenia solium".mp. or exp Taenia solium/ | 2,722 |
| 73 | "pork tapeworm".mp. | 201 |
| 74 | cysticercosis.mp. or exp Cysticercosis/ | 7,805 |
| 75 | 71 or 72 or 73 or 74 | 9,906 |
| 76 | trachoma.mp. or exp Trachoma/ | 5,417 |
| 77 | "Chlamydia trachomatis".mp. or exp Chlamydia trachomatis/ | 17,576 |
| 78 | 76 or 77 | 21,901 |
| 79 | exp Yaws/ or yaws.mp. | 1,217 |
| 80 | "Treponema pallidum pertenue".mp. | 13 |
| 81 | 79 or 80 | 1,221 |
| 82 | 4 or 7 or 11 or 15 or 16 or 26 or 30 or 33 or 36 or 39 or 44 or 45 or 49 or 50 or 54 or 57 or 67 or 70 or 75 or 78 or 81 | 329,331 |
| 83 | "political violence".mp. | 519 |
| 84 | demonstrations.mp. | 8,157 |
| 85 | "strategic developments".mp. | 44 |
| 86 | exp civil disorders/ or exp "warfare and armed conflicts"/ | 50,502 |
| 87 | exp Military Medicine/ | 30,241 |
| 88 | battle.mp. | 10,020 |
| 89 | exp "Dissent and Disputes"/ | 6,783 |
| 90 | protest.mp. | 1,771 |
| 91 | riot.mp. or exp Riots/ | 682 |
| 92 | exp Chemical Warfare/ or exp Chemical Warfare Agents/ or "chemical weapon".mp. | 30,236 |
| 93 | "drone strike".mp. | 1 |
| 94 | exp Terrorism/ | 13,923 |
| 95 | exp Weapons/ or weapons.mp. | 46,061 |
| 96 | exp human trafficking/ or exp rape/ or exp gender-based violence/ | 8,339 |
| 97 | "sexual violence".mp. | 5,889 |
| 98 | "forced disappearance".mp. | 10 |
| 99 | 83 or 84 or 85 or 86 or 87 or 88 or 89 or 90 or 91 or 92 or 93 or 94 or 95 or 96 or 97 or 98 | 162,057 |
| 100 | 82 and 99 | 856 |
| 101 | limit 100 to (english language and yr="2005 -Current") | 362 |

**Supplementary file 2: eligibility criteria in full**

Eligibility criteria

A PICOST framework was used to construct this review’s research question and eligibility criteria (1):

| Population/problem | NTD programmes in any conflict-related humanitarian emergency setting (as defined by ACLED codebook) |
| --- | --- |
| Intervention/exposure | Challenges encountered, and strategies or solutions implemented, by NTD programmes in CRHE to reaching NTD road map targets |
| Comparison | N/A |
| Outcome | Outcomes as reported by each study, including intervention acceptability and progress towards targets for each NTD as per WHO road map |
| Study Design | Primary research studies including observation studies, case reviews and randomised control trials. Opinion pieces and standalone cost-effectiveness and modelling studies excluded |
| Time Frame | Search limited to the years ACLED coding has been in place (1^st^ January 2005) until search date (30th May 2024) |

The criteria listed below were applied to all search results:

Inclusion criteria:

- Papers on humans
- Papers meeting ACLED codebook criteria for a conflict-related humanitarian emergency setting (2)
- Papers written from 1^st^ January 2005 (date when ACLED was founded (3)) to 30^th^ May 2024
- Papers on at least one WHO-defined NTD (4)
- Papers focused on NTD programmes in CRHE
- Papers discussing the challenges encountered by NTD programmes in conflict settings
- Papers written in English
- Papers reporting, or conducting analysis on, primary data

Exclusion criteria:

- Papers with missing abstract, full text, or other key information that cannot be found within the paper
- In circumstances of duplicate results, the duplicate results paper will be excluded (with preference for retaining the paper with final, as opposed to preliminary, results)
- Papers only using animal, cellular, genomic or laboratory models or data (non-humans)
- Papers not discussing interventions related to NTD programmes in CRHE
- Papers written in language other than English
- Secondary analysis papers, including mathematical modelling (due to results being a forecast of data) or systematic reviews (due to being a summary of analyses), and opinion articles (due to high-risk of bias or of being low-quality evidence)

**Supplementary file 3: Summary table of data extraction for 26 included papers**

| **Paper details**  **(First author, Title, Year)** | **Study description, including NTD focus and CRHE** | **Challenges encountered by NTD programme** | **Solutions / Strategies / Learnings reported in paper** | **Evidence gaps identified in paper** |
| --- | --- | --- | --- | --- |
| Hussein NR, A clinical study of cutaneous leishmaniasis in a new focus in the Kurdistan region, Iraq, 2019 (5) | Case series of CL in Iraq, from patients referred to regional health centre, which included IDPs due to civil conflict | - Collapse of health services, infrastructure including transport and living conditions during war  - Conflict led to break down of municipal services such as rubbish collection (breading ground of sandfly vector)  - Poor geographic access preventing full-case finding  - Population displacement spread leishmaniasis from endemic regions of Iraq  - Limited study funding hindered long-term follow-up or second-line treatment options | -Multisectoral activities needed to control vector, promote health education, increase community participation  - Study activities enabled more aggressive management which increased CL cure rates | 1) Unknown when is the optimum time in disease development to commence treatment  2) Poor investment in CL research or drug development |
| Turan E, A Comparison of Demographic and Clinical Characteristics of Syrian and Turkish Patients with Cutaneous Leishmaniasis, 2015 (6) | Retrospective cohort study of CL in Turkey, comparing clinical and demographic factors between Turkish patients to Syrian patients who fled Syrian Civil War as refugees into Turkey | - Movement of refugees spreading CL  - Delays accessing treatment  - Refugee camp living conditions increase potential contact with sandfly vector | Deeper epidemiological knowledge of CL required for control programmes | N/A |
| Zeng W, Associations among Soil-Transmitted Helminths, G6PD Deficiency and Asymptomatic Malaria Parasitemia, and Anemia in Schoolchildren from a Conflict Zone of Northeast Myanmar, 2020 (7) | Cross-sectional survey of school children in IDP-settlement and township in North-East Myanmar (constructed during civil conflict) comparing interactions between STH, malaria, G6PD-deficiency and their impact on anaemia. | - Mass population movement due to conflict  - Poor living conditions in IDP camps  - Minimal healthcare infrastructure and resources to control parasitic infections | STH and malaria control strategies need to integrate  micronutrient deficiencies and anaemia. | No trials on the impact of malaria MDA strategies in children also living with STH and anaemia |
| Ullah Z, Characterizing cutaneous leishmaniasis in a conflict-affected region: a study from North Waziristan, Pakistan, 2023 (8) | Observational prospective study on CL patients attending regional referral centre in North Waziristan, Pakistan. This area has seen mass population displacement including cross-border movement (from neighbouring Afghanistan) due to decades of political instability, and ongoing military operations causing infrastructure collapse | - Limited funding/resources and high security concern limited study group access to field.  - Poor health infrastructure restricts access to healthcare  - Population movement spreading leishmaniasis and other infectious diseases from high endemicity regions  - Inadequate vector-control methods | - CL control programmes need political stability, health infrastructure, vector control, parasite detection measures and awareness campaigns  - Advanced molecular-based assays recommended for leishmaniasis speciation  - CL distribution can be mapped using hospital records  - CL cases largely in rural regions and in people living in non-cemented houses | N/A |
| Semage SN, Cutaneous leishmaniasis in Mullaitivu, Sri Lanka: a missing endemic district in the leishmaniasis surveillance system, 2014 (9) | Descriptive case report analysis of CL collected through Sri Lanka Army national surveillance system, which operated during and after Sri Lankan civil war, and was kept separate from national reported statistics. | - Poor CL awareness, little diagnostic capacity and resources in public health system. Army system had adequate personnel and resources throughout the country, standardising disease surveillance  - Separate reporting systems for Army and population gives unreliable national-level data, likely gross underestimation of true disease burden | - Active surveillance of CL in high-prevalence regions such as resettlement camps for IDPs and soldiers | N/A |
| Muhjazi G, Cutaneous leishmaniasis in Syria: A review of available data during the war years: 2011–2018, 2019 (10) | Observational case reports of CL in Syria during civil war, which involved mass-population displacement. Country-level data extracted from 3 systems: 1) Ministry of Health surveillance 2) WHO EWARS 3) MENTOR initiative (NGO collected surveillance data) | - Poor coverage of surveillance systems, with limited healthcare workers and resources to provide diagnostics and treatment  - Unstable political situation gives unreliable data – stability sees a surge in cases due to increased detection  - Large-scale population movement affect epidemiology of disease (from endemic to non-endemic areas)  - Overcrowding and poor living conditions due to conflict | - Decrease in cases seen when government and international health organisations made efforts to improve healthcare access and distribute insecticide-treated nets  - Cessation of fighting key for CL control programmes. | N/A |
| Murugananthan K, Demographic and clinical features of suspected dengue and dengue haemorrhagic fever in the Northern Province of Sri Lanka, a region afflicted by an internal conflict for more than 30 years-a retrospective analysis, 2014 (11) | Retrospective analysis of dengue cases from a state teaching hospital in Northern province of Sri Lanka. Conflict in North Sri Lanka for two decades, ending in 2009 - during active conflict there was no movement into this region, but afterwards mass movement into and out of region | - Mass population movement at end of conflict, shifting disease epidemiology and overwhelmed an unprepared health and surveillance system (due to prior years of low prevalence)  - Poor education, awareness and clinical experience of community doctors with dengue fever leading to low levels of clinical suspicion – demonstrated by frequent delays in care and admissions late into symptom development.  - Lack of centralised reporting or case-notification system, alongside poor laboratory diagnostic capacity to confirm cases led to poor outcomes for patients, and poor knowledge of the scale of dengue burden | - Ministry of Health should oversee central reporting system for positive cases  - Appropriate training of community/primary healthcare professionals who have early contact with patients  - Increasing laboratory diagnostic capacity with regional state-sector laboratories having stock of antibody-based and virological tests to aid quicker diagnosis after clinical suspicion, enabling quicker hospital care to minimise complication risk | N/A |
| Alghazali KA, Dengue fever among febrile patients in Taiz City, Yemen during the 2016 war: Clinical manifestations, risk factors, and patients’ knowledge, attitudes, and practices toward the disease, 2019 (12) | Cross-sectional study of dengue in a hospital in Taiz city in South-West Yemen, region of intense conflict during civil war | - Ministry of Health had poor capacity for diagnostics and outbreak-response pre-conflict, which was further worsened by war. Widespread resource shortages, leaving reliance on clinical presentations to diagnose.  - War destroyed and disrupted infrastructure (road and water-supply damage), cessation of public services such as waste collection, plus military zones created around cities (involving creation of camps and trenches). This led to rubbish accumulation, open trenches and reliance on stored water (often in open containers) which are breeding grounds for mosquito vector, leading to higher risk of disease  -Difficulties obtaining and transporting patient samples in active conflict due to poor road access and security risk  - Conflict disrupted disease interventions (dengue awareness campaigns, distribution of mosquito nets and insecticides). Resulted in poor knowledge of dengue symptoms leading to delays in seeking healthcare, and minimal resources to distribute prevention measures  - Awareness of preventative measures did not translate into practice (little use of bed-nets or insecticide spraying. Ongoing war caused instability and mass-unemployment, so financial concerns meant individuals prioritised basic needs over dengue-preventative measures  - Increased refugees led to house overcrowding - men often staying outside houses, being exposed to mosquito bites thus increased risk of dengue  - Widespread famine and malnutrition in war, increasing susceptibility to infections | - Public health initiatives directly targeting population needed to increase education about clinical signs and risk factors of dengue  - Community participation should be prioritised in low-resourced health services which are often unable to provide detailed diagnostics | N/A |
| Inojosa WO, Diagnosing Human African Trypanosomiasis in Angola using a card agglutination test: observational study of active and passive case finding strategies, 2006 (13) | Observational clinical study of HAT based in Northern Angola during final stages of active civil war, comparing card agglutination test to conventional parasitological methods in active versus passive case finding techniques | - Conflict and instability challenged active case finding ability, preventing collection of repeated clinical samples (so lack of long-term follow-up and chance of not capturing positive rate accurately)  - Difficulty accessing all regions - often only single examination available for villagers (rather than repeated examinations which is WHO recommendation)  - Conflict severely inhibited resources and funding, so using only one test (card agglutination) attractive option. However, study showed this results in false-positive patients being subject to potentially harmful treatment | - Two-stage screening tests (albeit more resource intensive) are appropriate for accurate diagnostics and avoidance of inappropriate exposure to toxic therapies  - Once sufficient political stability achieved, HAT control programmes should transition from crisis intervention during humanitarian emergency programmes to more considered surveillance and containment programmes | N/A |
| Inci R, Effect of the Syrian Civil War on Prevalence of Cutaneous Leishmaniasis in Southeastern Anatolia, Turkey, 2015 (14) | Retrospective analysis of CL case records based in tertiary health centre in Anatolia, Turkey. Study population included refugees from neighbouring Syria who had fled civil war and now living in tent camps | - Changing CL epidemiology due to population movement - higher proportion of female than male patients due to female majority of refugees (males stayed in Syria or were casualties of war)  - Poor hygiene and housing in refugee camps facilitating spread of infectious diseases  - Difficulties in accessing healthcare in rural areas where camps situated | - Improvement of refugee housing conditions entering from a highly endemic area to prevent further spread of CL disease  - Effective CL case management must be in place to prevent infection spread  - Vector control measures should be implemented in regions receiving large numbers of people from highly endemic regions | N/A |
| Safi N, Evaluation of thermotherapy for the treatment of cutaneous leishmaniasis in Kabul, Afghanistan: a randomized controlled trial, 2012 (15) | Prospective randomised control trial comparing local thermotherapy to current standard of intralesional injection for CL. Study based in Kabul, Afghanistan during ongoing conflict which caused mass movement across the country, increase in refugee population, and disruption of infrastructure | - Conflict caused healthcare disruption, limited equipment and facilities all increasing CL risk  - Changing CL epidemiology: mass movement of individuals (from endemic to non-endemic regions), disruption of treatment and vector-control strategies led to disease spread in previous non-endemic areas  - CL is challenging to treatment - frequent relapses, high reinfection rates, and complex treatment planning decisions (including prior treatments, disease severity, co-infections and leishmania species) |  | Little data on CL cure rates leading to insufficient evidence to create appropriate guidelines |
| Jamal M, Exploring multi-level risk factors and post-war burdens of trachomatous trichiasis among displaced population in Raya Kobo districts, implication for urgent action, 2023 (16) | Community-based cross-sectional study mapping trachoma prevalence and risk factors after conflict (in Amhara and Afar regions of Ethiopia). Data collection in slums/villages in Raya Jobo district, occupied during the war, with destruction of many healthcare facilities and population displaced | - Mass-displacement of population gave rise to large camps, with overcrowded conditions, poor water supply, food insecurity and lack of WASH facilities  - Widespread healthcare destruction during conflict prevented patients accessing healthcare, particularly early care for ocular infections | - Community-level preventative factors for trachoma infection (face washing and latrine availability) should be scaled-up in high prevalence settings (such as displaced populations)  - Risk factors for active trachoma infection include age over 45 years and distance from clean water source. These high-risk groups require clinical intervention and scale-up of SAFE strategy | N/A |
| Badia-Rius X, Impact of conflict on the elimination targets of lymphatic filariasis, schistosomiasis and soil-transmitted helminths in Cabo Delgado province, Mozambique, 2024 (17) | Analysis of publicly available data from Cabo Delgado province in Mozambique, site of conflict and violence since 2017, to determine impact of conflict on NTD programmes and the challenges faced in achieving the WHO roadmap targets by 2030 | - Population displacement hindered drug administration programme  - Conflict limits access of NTD programmes to local populations to conduct surveys and education campaigns  - Conflict disrupting NTD programmes has implications for logistics and financing of future programmes due to re-emergence or spread of disease.  - IDPs are a high-risk group for communicable diseases, often living in overcrowded conditions with poor access to water, food, hygiene, healthcare and employment opportunities | - Stratified assessment methods accounting for levels of geographic access, conflict and population displacement should be used when planning NTD programmes  - Specialist guidelines with modified survey methodologies to accurately assess transmission are need for hard-to-reach populations or those with high IDP-rates or living in conflict  - NTD programmes should include a wide variety of stakeholders when planning response to complex emergencies, including from humanitarian agencies | 1) Little data to quantify the impact of conflict on NTD programme targets, which is needed to understand the scale of impact  2) Operational research from NTD programmes is not currently available, needed to evaluate effectiveness of current approaches |
| Chandrasena TG, Intestinal parasites and the growth status of internally displaced children in Sri Lanka, 2007 (18) | Cross-sectional survey of children based in a refugee camp (established for displaced people during the Sri Lankan civil war) estimating prevalence of intestinal parasitic infections and corresponding nutritional status | - Widespread undernutrition, poor sanitation and unsafe drinking water in refugee camps | - Relocation of IDPs to secure areas of Sri Lanka, with appropriate rehabilitation programmes whilst they are resettled  - Refugee camps run by the state should have appropriate safe food and water supplies, with broad-spectrum antihelmintics considered alongside sanitation and education campaigns | N/A |
| Rehman K, Leishmaniasis in Northern Syria during Civil War, 2018 (19) | Epidemiological study evaluating leishmaniasis services in Northern Syria during a time of political unrest and military action with subsequent population movement. Data collected as part of MENTOR initiative - a US and UK backed leishmania surveillance and control programme launched in 2013 | - No infrastructure in Syria to train and supply the healthcare teams required to implement and run leishmania programmes. Local workers often travelling to neighbouring Turkey to receive training and supplies which is dangerous  - No accurate population demographics – estimations from international institutions but there was lots of change due to war-related casualties and population displacement  - Fragmented healthcare system, and evolving security threats affected availability of healthcare facilities  - Conflict altered leishmania epidemiology due to: population movement, vector breeding in bomb-damaged buildings and lack of infrastructure leading to waste-accumulation in which vector can breed  -International Organisations offering CL aid during conflict used conflicting case definitions and reporting systems leading to unreliable data | - Leishmania control programmes depend upon political stability and security  - MENTOR initiative and its control programmes acted to avoid further increases in leishmania incidence seen after the onset of conflict. | Accurate population statistics lacking due to mass population movement, casualties of war |
| Dorkenoo, M.A., Monitoring migrant groups as a post-validation surveillance approach to contain the potential reemergence of lymphatic filariasis in Togo, 2021 (20) | Cross-sectional survey of 3 migrant groups in North Togo - one nomadic, one seasonal labour migrants and one refugees from Ghana (fleeing conflict, based in camps in Tandjouare district), to assess prevalence of LF infection and the risk posed by migrant groups to Togo's elimination of the disease | - Migration of people from endemic regions to non-endemic regions risk disease-spread and threaten elimination programmes  - Non-legal border crossing by migrants not included in official border surveillance programmes which threaten undocumented LF introduction  - Poor coverage of insecticide-treated nets (ITNs) reported, as distributions stopped once Togo deemed to have achieved LF elimination.  - Post achieving LF-elimination, LF programme funding rapidly decreased, so little resource to continue sustainable long-term surveillance | - Rapid post-validation surveys once LF programmes meet criteria of elimination as a public health problem are important, especially in settings like Togo where neighbouring countries are still LF-endemic. Surveys should be part of sustainable surveillance systems and integrated into wider health structure  - ITNs are an important prevention strategy, particularly in post-MDA regions to prevent disease re-introduction | Post-validation surveillance systems were absent in Togo |
| de Souza DK, No Evidence for Lymphatic Filariasis Transmission in Big Cities Affected by Conflict Related Rural-Urban Migration in Sierra Leone and Liberia, 2014 (21) | Cross-sectional survey to assess LF transmission in large cities in Liberia and Sierra Leone, to establish whether MDA is required for LF control. Surveys conducted after the end of Sierra Leone civil war (from 1991-2001), which saw an estimated 47% of pre-war population displaced either internally in camps or to neighbouring countries (Liberia, Guinea) | - Mass movement of population, largely from rural high-LF endemic regions to urban non-endemic areas | - Urban MDA programmes based on antigen prevalence alone are not reliable and can lead to over-treatment. Transient population movement due to conflict was not found to evidence LF transmission in these areas. Active transmission surveys measuring microfilaremia carrier prevalence at sentinel sites should be used instead | N/A |
| Suykerbuyk P, Persistence of Mycobacterium ulcerans disease (Buruli Ulcer) in the historical focus of Kasongo Territory, the Democratic Republic of Congo, 2009 (22) | Cross-sectional survey to identify prevalence and epidemiology of BU in Eastern DRC - site of conflict in 1990s (which spread to bordering Rwanda and Burundi). Mass population displacement and collapse of infrastructure, particularly healthcare due to security and geographic isolation of this region | - Poor living conditions (poor sanitation, access to healthcare, general infrastructure, transport) all increase BU risk  - Security risk, isolation and infrastructure breakdown during conflict meant large parts of the province became inaccessible to healthcare and aid workers  - Movement of people from BU-endemic regions to non-endemic areas | - Decentralise BU control programmes to community level for increased case management effectiveness  - Accessible soap and clean water, and ability to clean injuries in all communities required  - BU programmes should work alongside other community level control programmes existing for tuberculosis and leprosy | 1) Poor knowledge base of BU biology and clinical course  2) Little BU epidemiological data - particularly in historical foci in DRC |
| Alawieh A. Revisiting leishmaniasis in the time of war: the Syrian conflict and the Lebanese outbreak, 2014 (23) | Evaluation of leishmaniasis case reports from Ministry of Public Health surveillance department in Lebanon, to assess epidemiology post-influx of refugees from neighbouring Syria fleeing war, to aid control programmes and resource planning | - Influx of refugees from endemic Syria to non-endemic Lebanon, leading to increased leishmaniasis surveillance and reporting, so unclear whether true case increase or rather increased detection  - Densely-packed camps high-risk for leishmaniasis spread. But not all refugees entering camps –these people are missed by camp-targeted control efforts  - Rapid refugee movement did not allow healthcare system to prepare, and simultaneous increase of multiple infectious diseases occurred in a system already with little capacity  - Refugees delayed attending healthcare due to unfamiliarity with new country's healthcare system  - Limited diagnostic capacity due to lack of laboratory staff training  - Sparse healthcare coverage in rural and poor regions, where leishmaniasis predominantly affects, so unreliable surveillance data in these communities and extensive morbidity and stigma reported | - Role of media in education of leishmaniasis as war-related disease, helped to address stigma and encourage health-seeking behaviour  - Importance of speciating leishmaniasis to ensure appropriate treatment options  - Outbreaks should be addressed with a coordinated response from aid organisations and local and national governments | No vaccine and limited therapeutic options particularly in cases of resistant leishmania strains |
| Hastings J. Rumours, riots and the rejection of mass drug administration for the treatment of schistosomiasis in Morogoro Tanzania, 2016 (24) | Ethnography based on semi-structured interviews conducted in Morogoro region of Tanzania, where rioting began in 2008 after reports children died after receiving MDA at their primary school. Parents/Guardians stormed school buildings, teachers attacked, and school buildings damaged | - Deep-rooted concerns about MDA being linked to secret sterilisation campaigns by central government  - ‘Top down' approach to MDA programme - little community involvement, lack of acknowledgement for parental concerns - can spark violence and community discontent due to neglecting social and political contexts of those who have disease | - Social sciences and Anthropology have a place in planning NTD programmes, to ensure lived experiences of those with NTDs are not overlooked by programme designers  - To address parental concerns, MDA could be distributed by healthcare workers in hospitals/clinics, rather than schoolteachers | N/A |
| Whitehall JS, Snake bites in north east Sri Lanka, 2007 (25) | Retrospective case-review of patients with snake bites presenting to Kilinochchi regional hospital in North-East Sri Lanka - site of civil conflict for 20+ years, with regional economic sanctions resulting in resource shortage. Study conducted in 2005 during cease-fire period | - Shortage of healthcare workers and supplies impacted clinical care available for patients  - Unreliable drug supply, with antivenom available during study period because of cease-fire but not pre- or post-study during active fighting. Non-conflict zones reported antivenom cost double what it is in this conflict zone | - Good public education campaigns emphasising importance of early attendance to healthcare facilities led to low rate of complications  - Standardisation of therapy and access to antivenom and other essential medicines reduces mortality and morbidity across all ages for snake bites – such access is risked in active conflict and need to be protected  - Public health measures to address snake bites should include: removal of snake breeding and hiding sites near houses (1/4 of bites took place here), and advice for field workers to wear leggings and boots (nearly 4/5ths of bites occurred on feet or legs, particularly in harvest season) | N/A |
| Zhao Y, The epidemiology of 32 selected communicable diseases in Iraq, 2004-2016, 2019 (26) | Study of case reports to Ministry of Health (collected at primary health centres and public hospitals except Kurdistan region) to assess incidence of 32 communicable diseases during final decade of conflict in Iraq. Diseases divided into 4 categories: Cat 2 contains anthrax, rabies, visceral leishmaniasis; Cat 3 contains brucellosis, schistosomiasis, hydatidosis; Cat 4 contains leprosy, cutaneous leishmaniasis | - Lack of infrastructure investment due to UN sanctions on Iraq led to poor staffing, lack of drugs, medical supplies and equipment (specifically first-line treatments for leishmaniasis), and little capacity for disease treatment, surveillance or control  - Interruption to vaccination campaigns left population, particularly children, vulnerable to infectious disease  - Conflict caused population movement across country, facilitating disease spread and introduction of disease in non-immune populations. Displaced people often living in camps with poor food security and access to clean drinking water | - Enhanced vector control programmes and resumption of animal and vector programmes post- war led to declining / stable incidence of vector and zoonotic disease (brucellosis, schistosomiasis, hydatidosis, toxoplasmosis)  - Population given 'smart cards' which contain health records and allows reminders for when vaccinations are due  - Quick restoration and investment in surveillance systems and primary healthcare post-conflict ensured health system returned post war | Iraq’s Ministry of Health reports do not include private sector healthcare, any reports from Kurdistan region nor any data before the onset of war for comparison |
| Özkeklikçi A, The new situation of cutaneous leishmaniasis after Syrian civil war in Gaziantep city, Southeastern region of Turkey, 2017 (27) | Observational study of CL cases presenting to a state hospital in Gaziantep city, South-East Turkey, generating epidemiological knowledge via speciation diagnostic techniques. Region received many refugees fleeing civil war in neighbouring Syria, many of whom live in refugee camps | - Movement of people from Syria (region of high-endemicity) to Turkey (non-endemic) risks leishmaniasis spread  - Turkish government ‘Open Border’ policy to Syrian refugees means no checks when entering the country (so unknown whether disease being imported or transmitted within Turkey) and makes it hard to know specific numbers of those crossing or living in camps  - Lack of laboratory diagnostic equipment and capacity - only 34 of the 567 samples from CL-suspected patients underwent molecular diagnostics | - Vector control schemes need to be prioritised in regions where multiple leishmania species and vectors are detected as spreading | N/A |
| Macleod CK, Unimproved water sources and open defecation are associated with active trachoma in children in internally displaced persons camps in the Darfur States of Sudan, 2019 (28) | Secondary analysis of 27 cross-sectional population-based trachoma prevalence surveys, originally performed as part of Global Trachoma Mapping Project. Surveys covered 36 IDP camp clusters across 11 districts in Dafur states, West Sudan, site of war which started in 2003 ending in 2020, involving mass population displacement | - IDP camps are challenging environments (often crowded, some practice open defecation, water vendors can supply water in unimproved sources), all increasing trachoma infection risk (higher rates of trachoma in children in IDP camps than non-camps)  - Government-registered camps used for survey sites, potentially excluding more recent or non-official camps meaning true burden of disease may be unknown | - Water and sanitation access in IDP camps is challenging but necessary for trachoma control | 1) Poor evidence base and experience of SAFE strategy implementation in conflict settings such as IDP camps, with no guidelines for healthcare workers  2) Weak evidence base for WASH interventions during times of humanitarian crises |
| Yauba SM, Urinary schistosomiasis in Boko Haram-related internally displaced Nigerian children, 2018 (29) | Cross-sectional survey assessing prevalence and risk factors for urinary schistosomiasis in children living in IDP camps in Maiduguri, Nigeria. Camps were constructed to house displaced people fleeing violence and destruction of their homes due to rise of Boko Haram terrorist group, based in North-East Nigeria | - Changing schistosomiasis epidemiology affected programme planning – population displacement involves people from areas dependent on river water or farmers (high risk for schistosomiasis) moving to IDP camps (with a lower schistosomiasis burden) | - Community healthcare workers and paediatricians should visit local regions with high prevalence - particularly in areas housing lots of displaced people  - All communities dependent on river/stream water should be given praziquantel to address urinary schistosomiasis infection levels | N/A |
| Youssef A, Visceral and Cutaneous Leishmaniases in a City in Syria and the Effects of the Syrian Conflict, 2019 (30) | Observational analysis of leishmaniasis records (cutaneous and visceral) to assess for epidemiological change pre- and post- conflict in Latakia region in Syria which received a surge of displaced people from nearby cities during civil conflict | - Disruption of healthcare, pharmaceutical production and infrastructure as a direct result of conflict, resulted in very poor health access  - Widespread population displacement, often from highly endemic regions to non-endemic regions  - Population movement away from fighting in rural areas into cities led to urban overcrowding, poor conditions and increased infectious disease risk  - Poor medical records - hard to ascertain patient clinical histories, outcomes or specific treatments | - Concentrate living condition improvements to highly populous regions  - Early diagnostics, warning, control and education programmes vital for leishmaniasis programmes | Poor medical register, lack of details available on patients' clinical data during conflict times |

**Supplementary file 4: Summary of Joanna Briggs Institute (JBI) critical appraisal checklists for quality assessment of the 26 included papers.**

| **Paper Information**  (First author, Title, Year) | **Study type** | **JBI checklist** | **Score** [meets criteria / total applicable questions on checklist] (%) |
| --- | --- | --- | --- |
| Hussein NR, A clinical study of cutaneous leishmaniasis in a new focus in the Kurdistan region, Iraq, 2019 (5) | Case report analysis | Case series | 7/10 (70.0) |
| Turan E, A Comparison of Demographic and Clinical Characteristics of Syrian and Turkish Patients with Cutaneous Leishmaniasis, 2015 (6) | Retrospective cohort study | Case series | 5/8 (62.5) |
| Zeng W, Associations among Soil-Transmitted Helminths, G6PD Deficiency and Asymptomatic Malaria Parasitemia, and Anemia in Schoolchildren from a Conflict Zone of Northeast Myanmar, 2020 (7) | Cross-sectional survey | Analytical cross sectional | 6/8 (75.0) |
| Ullah Z, Characterizing cutaneous leishmaniasis in a conflict-affected region: a study from North Waziristan, Pakistan, 2023 (8) | Observational study | Case series | 9/9 (100) |
| Semage SN, Cutaneous leishmaniasis in Mullaitivu, Sri Lanka: a missing endemic district in the leishmaniasis surveillance system, 2014 (9) | Case report analysis | Case series | 6/7 (85.7) |
| Muhjazi G, Cutaneous leishmaniasis in Syria: A review of available data during the war years: 2011–2018, 2019 (10) | Case report analysis | Case series | 5/7 (71.4) |
| Murugananthan K, Demographic and clinical features of suspected dengue and dengue haemorrhagic fever in the Northern Province of Sri Lanka, a region afflicted by an internal conflict for more than 30 years-a retrospective analysis, 2014 (11) | Case report analysis | Case series | 6/8 (75.0) |
| Alghazali KA, Dengue fever among febrile patients in Taiz City, Yemen during the 2016 war: Clinical manifestations, risk factors, and patients’ knowledge, attitudes, and practices toward the disease, 2019 (12) | Cross-sectional survey | Analytical cross sectional | 6/6 (100) |
| Inojosa WO, Diagnosing Human African Trypanosomiasis in Angola using a card agglutination test: observational study of active and passive case finding strategies, 2006 (13) | Observational clinical study | Diagnostic Test Accuracy | 8/10 (80.0) |
| Inci R, Effect of the Syrian Civil War on Prevalence of Cutaneous Leishmaniasis in Southeastern Anatolia, Turkey, 2015 (14) | Case report analysis | Case series | 9/10 (90.0) |
| Safi N, Evaluation of thermotherapy for the treatment of cutaneous leishmaniasis in Kabul, Afghanistan: a randomized controlled trial, 2012 (15) | Prospective randomised controlled trial | Randomized Controlled Trials | 7/13 (53.8) |
| Jamal M, Exploring multi-level risk factors and post-war burdens of trachomatous trichiasis among displaced population in Raya Kobo districts, implication for urgent action, 2023 (16) | Cross-sectional survey | Analytical cross sectional | 8/8 (100) |
| Badia-Rius X, Impact of conflict on the elimination targets of lymphatic filariasis, schistosomiasis and soil-transmitted helminths in Cabo Delgado province, Mozambique, 2024 (17) | Case report analysis | Case series | 7/7 (100) |
| Chandrasena TG, Intestinal parasites and the growth status of internally displaced children in Sri Lanka, 2007 (18) | Cross-sectional survey | Analytical cross sectional | 6/8 (75.0) |
| Rehman K, Leishmaniasis in Northern Syria during Civil War, 2018 (19) | Epidemiological study | Case series | 10/10 (100) |
| Dorkenoo, M.A., Monitoring migrant groups as a post-validation surveillance approach to contain the potential reemergence of lymphatic filariasis in Togo, 2021 (20) | Cross-sectional survey | Analytical cross sectional | 8/8 (100) |
| de Souza DK, No Evidence for Lymphatic Filariasis Transmission in Big Cities Affected by Conflict Related Rural-Urban Migration in Sierra Leone and Liberia, 2014 (21) | Cross-sectional survey | Prevalence | 8/9 (88.9) |
| Suykerbuyk P, Persistence of Mycobacterium ulcerans disease (Buruli Ulcer) in the historical focus of Kasongo Territory, the Democratic Republic of Congo, 2009 (22) | Cross-sectional survey | Analytical cross sectional | 6/8 (75.0) |
| Alawieh A. Revisiting leishmaniasis in the time of war: the Syrian conflict and the Lebanese outbreak, 2014 (23) | Case report analysis | Case series | 10/10 (100) |
| Hastings J. Rumours, riots and the rejection of mass drug administration for the treatment of schistosomiasis in Morogoro Tanzania, 2016 (24) | Ethnography | Qualitative research | 9/10 (90.0) |
| Whitehall JS, Snake bites in north east Sri Lanka, 2007 (25) | Retrospective case review | Case series | 9/10 (90.0) |
| Zhao Y, The epidemiology of 32 selected communicable diseases in Iraq, 2004-2016, 2019 (26) | Case report study | Case series | 7/8 (87.5) |
| Özkeklikçi A, The new situation of cutaneous leishmaniasis after Syrian civil war in Gaziantep city, Southeastern region of Turkey, 2017 (27) | Observational study | Case series | 9/9 (100) |
| Macleod CK, Unimproved water sources and open defecation are associated with active trachoma in children in internally displaced persons camps in the Darfur States of Sudan, 2019 (28) | Secondary analysis of cross-sectional surveys | Analytical cross sectional | 8/8 (100) |
| Yauba SM, Urinary schistosomiasis in Boko Haram-related internally displaced Nigerian children, 2018 (29) | Cross-sectional survey | Prevalence | 8/9 (88.9) |
| Youssef A, Visceral and Cutaneous Leishmaniases in a City in Syria and the Effects of the Syrian Conflict, 2019 (30) | Observational analysis | Case series | 8/10 (80.0) |

2.	The Armed Conflict Location & Event Data Project (ACLED) ACLED Codebook, 2023. 2023.

3.	The Armed Conflict Location & Event Data Project (ACLED) ACLED History. 2022.

4.	World Health Organization Global report on neglected tropical diseases 2024. Geneva2024 [Available from: <https://www.who.int/publications/i/item/9789240091535> , date accessed 26.05.2025.

5.	Hussein NR, Balatay AA, Saleem ZSM, Hassan SM, Assafi MS, Sheikhan RS, et al. A clinical study of cutaneous leishmaniasis in a new focus in the Kurdistan region, Iraq. PLoS ONE. 2019;14(5).

6.	Turan E, Yeilova Y, Surucu HA, Ardic N, Doni N, Aksoy M, et al. A Comparison of Demographic and Clinical Characteristics of Syrian and Turkish Patients with Cutaneous Leishmaniasis. American Journal of Tropical Medicine and Hygiene. 2015;93(3):559-63.

7.	Zeng W, Malla P, Xu X, Pi L, Zhao L, He X, et al. Associations among Soil-Transmitted Helminths, G6PD Deficiency and Asymptomatic Malaria Parasitemia, and Anemia in Schoolchildren from a Conflict Zone of Northeast Myanmar. American Journal of Tropical Medicine & Hygiene.102(4):851-6.

8.	Ullah Z, Samad F, Bano R, Arif S, Zamir S, Aziz N, et al. Characterizing cutaneous leishmaniasis in a conflict-affected region: a study from North Waziristan, Pakistan. Turkish Journal of Medical Sciences.53(6):1767-75.

9.	Semage SN, Pathirana KPN, Agampodi SB. Cutaneous leishmaniasis in Mullaitivu, Sri Lanka: A missing endemic district in the leishmaniasis surveillance system. International Journal of Infectious Diseases. 2014;25:53-5.

10.	Muhjazi G, Gabrielli AF, Ruiz-Postigo JA, Atta H, Osman M, Bashour H, et al. Cutaneous leishmaniasis in Syria: A review of available data during the war years: 2011-2018. PLoS Neglected Tropical Diseases.13(12):e0007827.

11.	Murugananthan K, Kandasamy M, Rajeshkannan N, Noordeen F. Demographic and clinical features of suspected dengue and dengue haemorrhagic fever in the Northern Province of Sri Lanka, a region afflicted by an internal conflict for more than 30 years-a retrospective analysis. International Journal of Infectious Diseases.27:32-6.

21.	de Souza DK, Sesay S, Moore MG, Ansumana R, Narh CA, Kollie K, et al. No evidence for lymphatic filariasis transmission in big cities affected by conflict related rural-urban migration in Sierra Leone and Liberia. PLoS Neglected Tropical Diseases.8(2):e2700.
